# Absence of Testicular Adrenal Rest Tumors in Newborns, Infants, and Toddlers with Classical Congenital Adrenal Hyperplasia

**DOI:** 10.1101/19005975

**Authors:** Mimi S. Kim, Christina M. Koppin, Pankhuri Mohan, Fariba Goodarzian, Heather M. Ross, Mitchell E. Geffner, Roger De Filippo, Paul Kokorowski

**Author notes:** Corresponding Author: Mimi Kim, M.D., M.Sc., Children’s Hospital Los Angeles, 4650 Sunset Boulevard, Mailstop #61, Los Angeles, CA 90027, E-mail address.

## Abstract

**INTRODUCTION:** Testicular adrenal rest tumors (TART) are a known consequence for males with classical congenital adrenal hyperplasia (CAH) due to 21-hydroxylase deficiency. TART are associated with potential infertility in adults. However, little is known about TART in very young males with CAH.

**OBJECTIVE:** We assessed the prevalence of TART in newborn, infant, and toddler males with classical CAH via scrotal ultrasound.

**METHODS:** Males with CAH had scrotal ultrasounds during the first 4 years of life, evaluating testes for morphology, blood flow, and presence of TART. Newborn screen 17-hydroxyprogesterone (17-OHP) and serum 17-OHP at the time of ultrasound were recorded. Bone ages were considered very advanced if ≥ 2SD above chronological age.

**RESULTS:** Thirty-one ultrasounds in 16 males were performed. An initial ultrasound was obtained in four newborns at diagnosis (6.8 ±2.1 days), six infants (2.2 ±0.9 months), and six toddlers (2.4 ±0.9 years). Eleven males had at least one repeat ultrasound. A large proportion (11/16) were in poor hormonal control with an elevated 17-OHP (325 ±298 nmol/L). One infant was in very poor hormonal control (17-OHP 447 nmol/L) at initial ultrasound, and two toddlers had advanced bone ages (+3.2 and +4.5 SD) representing exposure to postnatal androgens. However, no TART were detected in any subjects.

**CONCLUSIONS:** TART were not found in males up to 4 years of age with classical CAH despite settings with expected high ACTH drive. Further research into the occurrence of TART in CAH may elucidate factors which contribute to the detection and individual predisposition to TART.

## Introduction

Classical congenital adrenal hyperplasia (CAH) due to 21-hydroxylase deficiency is a potentially life-threatening genetic cause of primary adrenal insufficiency characterized by cortisol, aldosterone, and epinephrine deficiencies, as well as androgen excess. It is an autosomal recessive disorder, characterized by a mutation in the *CYP21A2* gene which encodes the steroid 21-hydroxylase enzyme needed for the biosynthesis of cortisol and aldosterone [1, 2]. Since the production of cortisol is hindered in CAH due to 21-hydroxylase deficiency, there is a loss of negative feedback to the hypothalamus/pituitary gland and subsequent ACTH hyperstimulation ensues. These high levels of ACTH in CAH patients have been shown to have deleterious effects and may contribute to the development of testicular adrenal rest tumors (TART).

In an individual who develops TART, one would expect the ectopic adrenal tissue to be present from the earliest moments of gonadal development [3, 4]. Testicular adrenal rest cells originate from adrenal cells which migrate with primitive gonadal cells from the urogenital ridge at approximately the eighth week of fetal development [5]. As a result, adrenal rest cells in males may come to reside within the testicles in prenatal life and ultimately reside anywhere along the path of their descent [6].

TART are a well-known potential complication of CAH in males. They can become large testicular masses causing compressive irreversible damage to the testicular parenchyma, ultimately leading to fibrosis, decreased spermatogenesis, and decreased germ cells as seen on biopsy specimens of patients [7, 8]. Localizing the position of the tumors and determining the size may help determine a fertility prognosis. Patients with TART also have higher concentrations of steroid production including 17-hydroxyprogesterone and androstenedione [9]. Identifying the tumor early can allow physicians to focus on treatment regimens to prevent infertility and control potential ectopic steroid production.

TART have been reported in adults and children as young as 4.1 years old by ultrasound, with a prevalence of 44% in adults and 21-33% in childhood [7, 10, 11]. At our center, TART have been diagnosed in males as young as 6 years old. TART was detected in 1/9 (11%) of patients 5-12 years old, and 4/19 (21%) of patients older than 12 years old [12]. Historically, we know that histological evidence of TART can be found in post-mortem infants as young as 8 weeks old [3]. However, there is little known about the prevalence of TART in male newborns, infants, or toddlers with CAH.

Therefore, the aim of our study was to determine the prevalence of TART by scrotal ultrasound in newborn, infant, and toddler males with CAH due to 21-hydroxylase deficiency at a tertiary referral center for CAH. We hypothesized that TART would only be found in those with extremely poor hormonal control in this very young age group.

## Participants and Methods

### Study Participants

Subjects were recruited from our tertiary referral center for CAH. Inclusion criteria consisted of male sex with classical CAH due to 21-hydroxylase deficiency confirmed by biochemical and/or genetic testing, including newborns (< 1 month old), infants (1 to 12 months old), and toddlers (older than 12 months up to 4 years old). Exclusion criteria consisted of female sex, CAH not due to 21-hydroxylase deficiency, or age ≥4 years.

### Study Design

Scrotal ultrasounds (14-MHz linear array transducer) were performed prior to the age of 4 years. Testes were evaluated for morphology and blood flow, with particular emphasis on examining for the presence of lesions consistent with TART. Newborn screening for CAH, via elevations in 17-hydroxyprogesterone (17-OHP), is state mandated in California and has an estimated sensitivity of 98.5% and specificity of 99.9%, with activation cut-offs stratified by weight and gestational age. Newborn screening 17-OHP levels were recorded as a measure of disease severity at birth, and serum 17-OHP and androgen (testosterone and androstenedione) levels closest to time of imaging were recorded as measures of hormonal control.

The most recent hydrocortisone replacement dose (mg/m2/day) each subject maintained for at least three weeks prior to the time of ultrasound was recorded. Hydrocortisone replacement dose was not recorded for subjects without a stable dose for at least three weeks prior to the time of ultrasound.

Bone age was assessed in subjects when there was clinical concern, using a left hand x-ray to examine for signs of epiphyseal maturation, and read by a pediatric endocrinologist (M.S.K.). Bone age was not routinely performed in our young cohort of infants and toddlers. A bone age more than two standard deviations from chronological age were considered to be very advanced.

Descriptive statistics are presented as mean ± SD.

## Results

### Study Population

Sixteen males with CAH due to 21-hydroxylase deficiency were studied. Fifteen males had the salt-wasting form and one had the simple-virilizing form. At the time of ultrasound, average hydrocortisone doses were 29.5 ± 11.4 mg/m2/day for infants less than six months, 17.1 ± 7.4 mg/m2/day for infants greater than six months, and 13.0 ± 4.1 mg/m2/day for toddlers. The average newborn screen 17-OHP for all participants was 424 ± 206 nmol/L (range 73 – 800).

### Scrotal Ultrasound

Thirty one ultrasounds were performed on 16 males. Eleven of 16 males had at least one repeat ultrasound up until 4 years of life. Four newborns had an initial ultrasound at CAH diagnosis; their mean newborn screen 17-OHP was 395 ± 180 nmol/L. Six infants (age 1 to 12 months old) had an initial ultrasound prior to 4 months of age. One male infant was in very poor hormonal control in (17-OHP was 447 nmol/L at the time of the initial ultrasound), but remained TART-negative on two additional ultrasounds at 7.5 and 13.6 months of age despite having an increase in 17-OHP to 1,025 nmol/L, and very advanced bone age (+3.2 SD). Six toddlers had their initial ultrasound betweenn 1 and 4 years of age. One toddler also had a very advanced bone age (+4.5 SD), likely representing substantial exposure to postnatal androgens, but did not exhibit TART on ultrasound. At the time of the ultrasounds, a large proportion of males were in poor hormonal control, with 11 of 16 males exhibiting a considerably elevated 17-OHP of 325 ± 298 nmol/L despite being on high doses of hydrocortisone in most cases.

Overall, no TART were detected on any scrotal ultrasounds. Normal testicular morphology and blood flow were noted in all subjects.

## Discussion/Conclusion

In this study we looked for the presence of TART in males as young as newborns with repeated ultrasounds throughout the first four years of life. No TART were detected by ultrasound in males with classical CAH up to 4 years of life, including newborns at the time of diagnosis, infants, and toddlers. Despite high serum 17-OHP levels at birth and poor hormonal control in 11 of 16 subjects, including two toddlers with very advanced bone ages, ultrasound screening for TART remained negative in our cohort. Although we did not measure plasma ACTH in the study subjects, it can be inferred that plasma concentrations would be high given the magnitude of the 17-OHP concentrations in many of the boys. Our results suggest that poor hormonal control, or high ACTH levels alone, do not seem to be sufficient to drive the growth of detectable adrenal rest tumors in this age group [11]. However, our negative findings could also simply be a result of an absence of adrenal rest tissue.

The prevalence of TART in males has been estimated to be 33% in boys and 44% in men with classical CAH [10, 13]. Although the prevalence of TART is lower in pediatric patients, several investigators have noticed the presence of TART in males as young as 4.1 years, 6.2 years, and 7.5 years of age [7, 10, 14]. Futhermore, newborns with CAH have significantly lower cortisol concentrations with compensatory elevation of plasma ACTH levels, which should theoretically promote growth of adrenal rest tissue and lead to development of TART [3, 15]. ACTH levels have been previously correlated to the prevalence of TART in adult males [16]. One possible explanation for the decreased prevalence of TART in pediatric patients, however, is that a prolonged postnatal duration of exposure to elevated ACTH levels is important in stimulating the growth of TART [17]. The role of ACTH is supported by the finding that early-stage TART can regress with high-dose glucocorticoid therapy. Furthermore, TART can be present in males with poorly controlled Addison disease [18] and in the ovaries and/or para-ovarian regions in females with poorly controlled CAH [19]. Further research on the presence of TART in ectopic ACTH syndromes (small cell lung cancers, islet cell tumors, etc.) could reinforce the theory that ACTH influences TART theory development.

The importance of early detection of TART has several advantages. First, the prevalence of TART in a newborn or infant could indicate a high ACTH concentration *in utero*, which could provide a CAH disease severity measure. Secondly, TART are a well-known comorbidity in males with CAH and can cause infertility and severe pain due to the compression of the testicular parenchyma. Patients with TART are counseled to consider infertility as a possible consequence and cryopreservation of sperm may be recommended to these patients in the earlier stages of tumor development. While these are consequences of long-standing TART, early identification may aid in addressing the tumor at a preliminary stage as several case studies have reported the use of intensified glucocorticoid treatments to prevent growth and promote regression of these tumors [9].

Autopsy studies suggest that the prevalence of adrenal rest cells is variable. One of the earliest reports suggests that ectopic adrenal tissue can be found on autopsy in 50% of healthy neonates and children [20]. Other autopsy studies have found aberrant adrenal tissue in 7.5% of individuals as an ovoid mass near the testes, with 1.5% located close to the rete testes [4]. Furthermore, ectopic adrenal tissue during exploration for hernia repairs has been noted in 2.7% of cases [21]. A study of 89 deceased male neonates, without CAH and specifically examining the testes, found that only 4 out of 115 (3.5%) contained TART and only one of these near the rete testis [7]. By contrast, another study found adrenal nodules on autopsy in the testes of 43% (3/7) of male CAH infants less than eight weeks of age and 100% (n=14) children less than 14 months of age [3]. This seems to suggest that while adrenal tissue is uncommonly found in the testes, there could be undetectable progenitor cells that require further stimulus to be able to grow to detectable levels. Hence the prevalence and natural course of regression of testicular adrenal rest tissue in infants with CAH merits further research and could yield an increased understanding of the mechanisms that promote tumor formation in some but not others. Overall, a more in-depth understanding of TART in male infants and toddlers with and without CAH could help inform future clinical screening guidelines.

Our study results should be interpreted in light of certain limitations. It is unclear whether the lack of detection of TART in our cohort is related to the limited sensitivity of ultrasonography or if TART are simply absent in these patients. Furthermore, our sample size is small with limited power to detect a small prevalence of TART. A larger cohort of very young males with CAH could allow us to better characterize the absence or presumed very low prevalence of TART. Finally, temporal variation in hormonal control cannot be captured with our available serum test results and could result in misclassification of those in good versus poor control.

In conclusion, TART can be a clinically significant co-morbidity in males with CAH. In our study, TART were not detected by ultrasonography in very young males up to 4 years of age with classical CAH, despite conditions of high ACTH drive at birth and pre-treatment, and conditions of poor hormonal control while on treatment. Further study is needed to understand individual predisposition to TART, the regression of testicular adrenal rest cells in infants, and the sensitivity of imaging modalities in the detection of TART.

## Data Availability

Data is available under IRB approved data-sharing with the PI.

## Acknowledgments

We gratefully thank the patients and families at our center for their participation, Janet Guerrero (CAH Clinic Coordinator), and CARES Foundation for support of the CAH Comprehensive Care Center at Children’s Hospital Los Angeles.

## Statements

### Statement of Ethics

The study was reviewed and approved by the Children’s Hospital Los Angeles institutional review board (CCI-13-00239 TART and CAH; CCI-12-0020 Natural History Study of CAH from Infancy). Parents gave written informed consent in accordance with the World Medical Association Declaration of Helsinki.

### Author Contributions

Mimi S. Kim, Paul Kokorowski, and Mitchell E. Geffner conceptualized and designed the study. Fariba Goodarzian performed analysis of ultrasound data and Christina M. Koppin, Pankhuri Mohan, and Heather M. Ross collected the data. Mimi S. Kim and Christina M. Koppin drafted the initial manuscript. All authors critically reviewed the manuscript and approved of the final manuscript as submitted.

---

**Table 1.** Scrotal Ultrasounds in Newborns, Infants, and Toddlers with Classical Congenital Adrenal Hyperplasia.

| CAH Type | Newborn 17-OHP (nmol/L) | Age at ultrasound (months) | 17-OHP at ultrasound (nmol/L) | Hydrocortisone dose at ultrasound (mg/m<sup>2</sup>/day) | Advanced Bone Age (> 2 SD) |
| --- | --- | --- | --- | --- | --- |
| SV | 73 | US1: 25.5 | US1: 73 | US1: 10.9 | N/A |
| SW | 170 | US1: 0.3<br>US2: 18.5 | US 1: 272<br>US2: 188 | US1: Newborn<br>US2: 18.2 | N/A |
| SW | 183 | US1: 1.5<br>US2: 22.5<br>US3: 35.4 | US1: 2.0<br>US2: 1.8<br>US3: 135 | US1: 19.0<br>US2: 11.7<br>US3: 11.8 | N/A |
| SW | 277 | US1: 3.7<br>US2: 7.5<br>US3: 13.6 | US1: 447<br>US2: 9.7<br>US3: 1025 | US1: 19.4<br>US2: 25.6<br>US3: 21.7 | Yes |
| SW | 281 | US1: 25.2<br>US2: 43.5 | US1: 39<br>US2: 43 | US1: 13.9<br>US2: 13.5 | No |
| SW | 340 | US1: 0.2 | US 1: 163 | US1: Newborn | N/A |
| SW | 363 | US1: 46.4 | US1: 1.4 | US1: 20.3 | Yes |
| SW | 394 | US1: 0.3<br>US2: 10.4 | US1: 868<br>US2: 3.8 | US1: Newborn<br>US2: 13.2 | N/A |
| SW | 406 | US1: 0.2<br>US2: 11.0 | US1: 663<br>US2: 2.2 | US1: Newborn<br>US2: 11.6 | N/A |
| SW | 492 | US1: 26.8<br>US2: 33.3 | US1: 4.0<br>US2: 104 | US1: 6.3<br>US2: 10 | No |
| SW | 502 | US1: 31.6 | US1: 15 | US1: 9.8 | N/A |
| SW | 550 | US1: 14.2<br>US2: 46.4 | US1: 28<br>US2: 150 | US1: 17.9<br>US2: 11.1 | No |
| SW | 611 | US1: 1.0<br>US2: 17.2 | US 1: 48<br>US2: 264 | US1: 40.0<br>US2: 13.0 | N/A |
| SW | 631 | US1: 1.8 | US1: 1.3 | US1: 41.7 | N/A |
| SW | 712 | US1: 1.5<br>US2: 23.9 | US1: 33<br>US2 : <0.2 | US1: 19.0<br>US2: 9.0 | N/A |
| SW | 808 | US1: 2.4<br>US2: 23.3<br>US3: 33.3 | US1: 4.1<br>US2: 274<br>US3: 205 | US1: 38<br>US2: 11.6<br>US3: 13.9 | No |

17-OHP, 17-hydroxyprogesterone; CAH, congenital adrenal hyperplasia; SV, simple-virilizing; SW, salt-wasting Hydrocortisone dose: At the time of ultrasound, subject was on the dose for less than three weeks. Newborns were not on a daily dose of hydrocortisone.

